# Impact of the population age, testing and vaccination levels on the numbers of COVID-19 cases and deaths per capita and case fatality risks

**DOI:** 10.1101/2023.12.09.23299758

**Authors:** Igor Nesteruk

## Abstract

The population, governments and researchers show much less interest in the COVID-19 pandemic. However, many questions still need to be answered: why the much less vaccinated African continent has accumulated 15 times less deaths per capita than Europe? or why in 2023 the global value of the case fatality risk is almost twice higher than in 2022 and the UK figure is four times higher than the global one?

The averaged daily numbers of cases *DCC* and death *DDC* per million, case fatality risks *DDC/DCC* were calculated for 34 countries and regions with the use of John Hopkins University (JHU) datasets and possible linear and non-linear correlations with the averaged daily numbers of tests per thousand *DTC*, median age of population *A*, and percentages of vaccinations *VC* and boosters *BC* were investigated. Strong correlations between age and *DCC* and *DDC* values were revealed. One-year increment in the median age yielded 39.8 increase in *DCC* values and 0.0799 DDC increase in 2022 (in 2023 these figures are 5.8 and 0.0263, respectively). With decreasing of testing level *DTC*, the case fatality risk can increase drastically. *DCC* and *DDC* values increase with increasing the percentages of fully vaccinated people and boosters, which definitely increase for greater *A*. After removing the influence of age, no correlations between vaccinations and *DCC* and *DDC* values were revealed.

The presented analysis demonstrates that age is a pivot factor of visible (registered) part of the COVID-19 pandemic dynamics. Much younger Africa has registered less numbers of cases and death per capita due to many unregistered asymptomatic patients. Of great concern is the fact that COVID-19 mortality in 2023 in the UK is still at least 4 times higher than the global value caused by seasonal flu.

## Introduction

In the fourth year of the COVID-19 pandemic, the population and governments show much less interest in it. In particular, only 39% of countries reported at least one case to WHO in the period from 31 July to 27 August 2023, [1]. Thus accumulated numbers of cases *CC* and deaths *DC* per million show stabilization trends [2] and can be used to estimate the impact of different factors on the pandemic dynamics and to answer some important questions. In particular, why the much less vaccinated African continent has accumulated 36 times less cases and 15 times less deaths per capita than Europe (see [1, 2], lines 26 and 27 in Table 1)? Why in 2023 the global value of the case fatality risk is almost twice higher than in 2022 and the UK figure is four times higher than the global one, [3])?

**Table 1.**
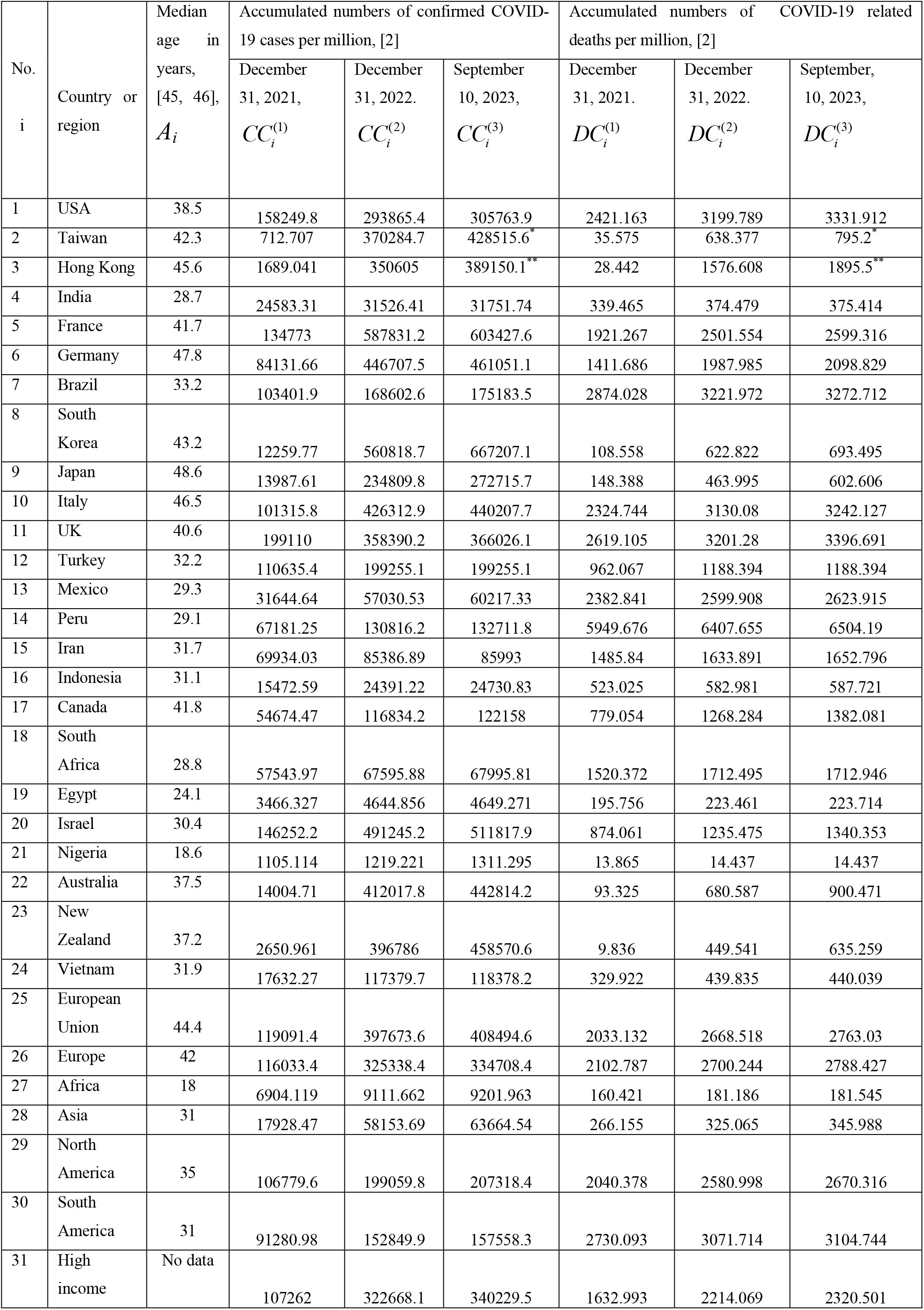
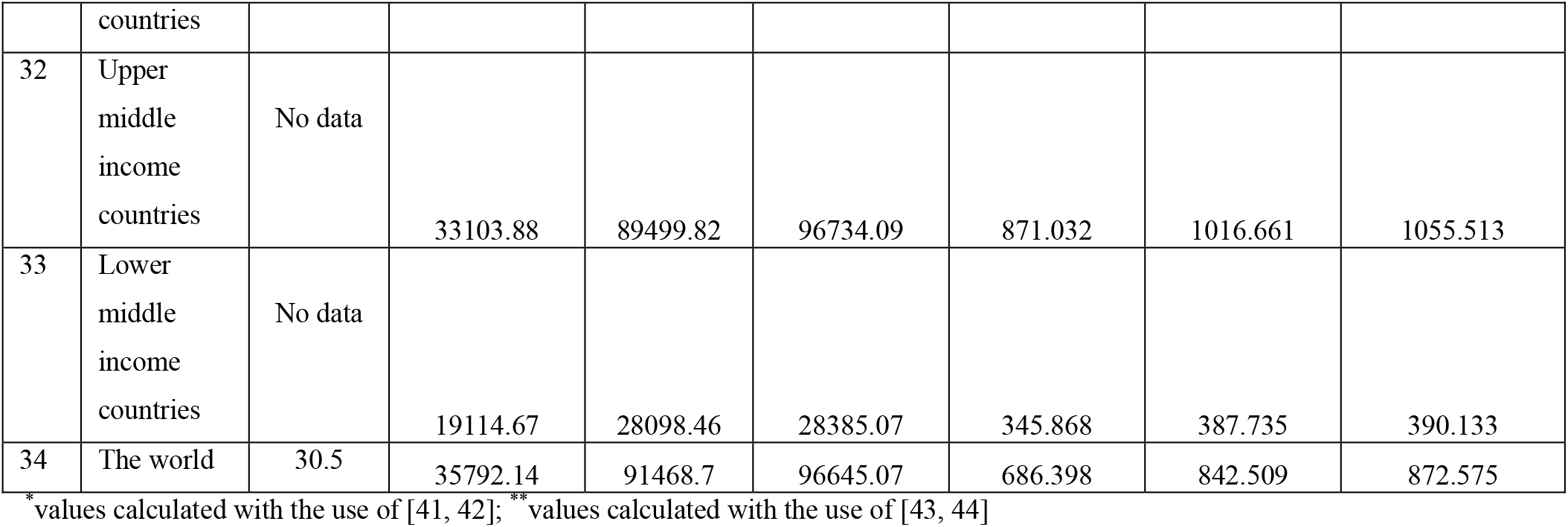
Median age, accumulated numbers the COVID-19 cases and deaths per capita in 2021-2023.

We will apply the linear correlation analysis using accumulated relative characteristics: the numbers of cases and deaths per million (*CC* and *DC*), numbers of fully vaccinated people and boosters per hundred (*VC* and BC), tests per thousand (*TC*) available in files of John Hopkins University (JHU) [2] for all countries and regions.

The impact of various factors on the COVID-19 pandemic dynamics was estimated in many papers. Some examples can be found in [4-21]. In particular, *CC* values accumulated as of December 23, 2021 in Ukrainian regions and European countries showed no correlations with the size of population, its density, and the urbanization level, while *DC* and *CFR=DC/CC* values reduce with the increase of the urbanization level in European countries, [18]. The increase of income (Gross Domestic Product per capita) leads to increase in *CC, VC, BC*, and *TC* values, but *DC* and *CFR* demonstrate opposite trend in European countries [19].

Many asymptomatic COVID-19 cases [22 - 26] can cause a big difference between visible (registered) and real pandemic dynamics [15, 27, 28]. That is why the higher testing level can increase the numbers of registered cases. Sometimes the testing level is too low to reveal all the cases predicted by theory. Probably such situation occurred in Japan in summer 2022, [29]. It was shown in [20], that the test per case ratio *TC/CC* (or test positivity rate *CC/TC*) is very important characteristic to control the pandemic. Very strong correlation between *CC* and *TC* was revealed for values accumulated before August 1, 2022 in European and African countries, [19]. Since the *TC* values have stopped to be updated by JHU in different days of 2022, [2, 19], in this study, we will investigate a correlation between the averaged daily numbers of cases and test per capita *DCC* and *DTC*, respectively.

The severity of SARS-CoV-2 infection increases for older patients [16]; almost half of the infected children can be asymptomatic [30]. With the use of the statistical analysis of 2020 datasets it was shown that younger populations have less clinical cases per capita and it was predicted that “without effective control measures, regions with relatively older populations could see disproportionally more cases of COVID-19, particularly in the later stages of an unmitigated epidemic” [31]. This forecast was confirmed in [21] with the use of *CC* and *DC* datasets for 79 countries and regions including 10 so-called Zero-COVID countries [32]. It was shown that one-year increment in the median age yields 12,000-18,000 increase in *CC* values and 52-83 increase in *DC* values. To confirm the same trends in 2022 and 2023, we will investigate correlations between median age *A* and the averaged daily numbers of cases and deaths per capita *DCC* and *DDC*, respectively.

The high numbers of circulating SARS-CoV-2 variants [33-35] and re-infected persons [36-38] raise questions about the effectiveness of vaccinations. In particular, many scientists are inclined to think that the pandemic will not be stopped only through vaccination [39]. The non-linear correlations show that *CC* and *DC* values increase with the growth of the vaccination level *VC*, while *CFR* decreases, [19]. In this study, we will investigate correlations between *VC* and *BC* values registered in 2022 and 2023 and *DCC, DDC* and *CFR* figures. We will try also to answer the question why the number of cases and death per capita are higher in more vaccinated countries.

## Materials and Methods

We will use the accumulated numbers of laboratory-confirmed COVID-19 cases *CC*_*i*_ and deaths *DC*_*i*_ per million, accumulated numbers of tests per thousand *TC*_*i*_, accumulated numbers of fully vaccinated people *VC*_*i*_ and boosters *BC*_*i*_ per hundred for 33 countries (shown in Tables 1 and 2) and the world (*i* =1, 2,.., 34). The have chosen the countries with highest numbers of accumulated cases and death (according to the recent WHO reports [1]), some other countries and regions listed in COVID-19 Data Repository by the Center for Systems Science and Engineering (CSSE) at Johns Hopkins University (JHU), [2] (version of file updated on September 28, 2023). Since Chinese statistics shows some contradictions (see, e.g., [40] or compare JHU files updated on September 28 and March 9, 2023), we have used only figures for Taiwan and Hong Kong. In particular, in Table 1 we show corresponding *CC*_*i*_ and *DC*_*i*_ values from the March-9-version of JHU file (not available on September 28). The *CC*_*i*_ and *DC*_*i*_ values for Taiwan and Hong Kong for 2023 were calculated with the use of [41-44]. We ignore data from Ukraine and Russia to exclude the influence of military operations on the COVID-19 statistics. Nevertheless, the aggregated figures for large regions, continents and the world include information from these two countries and China ( see lines 25-34 in Tables 1 and 2). To take into account the average age of population, we have used the information about the median ages *A*_*i*_ from [45, 46] (see Table 1).

**Table 2.** Accumulated numbers the tests per capita and percentage of fully vaccinated people and boosters in 2021-2023, [2].

| No., I | Country or region | Accumulated numbers of tests per thousand $TC_i^{(j)}$,<br>corresponding dates and number of days $T_i^{(TC)}$ | | | | | Numbers of fully vaccinated people<br>per hundred as of: | | Numbers of boosters<br>per hundred as of: | |
| --- | --- | --- | --- | --- | --- | --- | --- | --- | --- | --- |
| | | $TC_i^{(1)}$ | Date in 2021 | $TC_i^{(2)}$ | Date in 2022 | $T_i^{(TC)}$,<br>days | July 1. 2022<br>$VC_i^{(1)}$ | July 1. 2023<br>$VC_i^{(2)}$ | July 1. 2022<br>$BC_i^{(1)}$ | July 1. 2023<br>$BC_i^{(2)}$ |
| 1 | USA | 2155.384 | Dec 31 | 2708.533 | Jun 18 | 169 | 67.33 | 69.47<sup>23</sup> | 37.91 | 40.08<sup>8</sup> |
| 2 | Taiwan | 208.248 | Dec 31 | 545.749 | Jun 22 | 173 | 81.52 | 87 | 72.89 | 106.4 |
| 3 | Hong Kong | 4615.889 | Dec 31 | 6594.993 | May 24 | 144 | 86.21 | 90.81 | 60.36 | 94.91 |
| 4 | India | 481.597 | Dec 31 | 609.938 | Jun 21 | 172 | 64.58 | 67.17 | 3.15 | 16.04 |
| 5 | France | 2908.831 | Dec 31 | 4126.754 | Jun 18 | 169 | 78.14 | 78.44 | 58.86 | 70.34 |
| 6 | Germany | 1108.669 | Dec 26 | 1574.021 | Jun 12 | 168 | 76.04 | 76.24<sup>20</sup> | 68.6 | 77.72<sup>20</sup> |
| 7 | Brazil | 308.411 | Dec 31 | 330.912 | March 11 | 70 | 78.52 | 81.82<sup>17</sup> | 49.7 | 58.7<sup>17</sup> |
| 8 | South Korea | 874.47 | Dec 31 | 1934.578 | Jun 15 | 166 | 85.48 | 85.64<sup>25</sup> | 73.01 | 79.76<sup>14</sup> |
| 9 | Japan | 224.641 | Dec 31 | 429.37 | Jun 22 | 173 | 82.6 | 83.4<sup>22</sup> | 64.04 | 141.72<sup>22</sup> |
| 10 | Italy | 2365.578 | Dec 31 | 3795.998 | Jun 22 | 173 | 81.14 | 81.22 | 69.66 | 80.88 |
| 11 | UK | 5845.841 | Dec 31 | 7480.121 | May 19 | 139 | 74.36 | 75.19<sup>10</sup> | 59.26 | 59.81<sup>9</sup> |
| 12 | Turkey | 1403.53 | Dec 31 | 1924.668 | May 31 | 151 | 62.21<sup>6</sup> | 62.31<sup>13</sup> | 43.22<sup>6</sup> | 48.54<sup>13</sup> |
| 13 | Mexico | 95.052 | Dec 31 | 122.879 | Jun 18 | 169 | 62.7<sup>3</sup> | 64.19<sup>11</sup> | 41.65<sup>3</sup> | 44.73<sup>12</sup> |
| 14 | Peru | 646.299 | Dec 31 | 859.283 | April 05 | 95 | 81.44 | 84.21 | 61.41 | 90.41 |
| 15 | Iran | 477.494 | Dec 31 | 594.485 | Jun 01 | 152 | 65.34<sup>2</sup> | 66.15<sup>29</sup> | 31.11<sup>2</sup> | 32.27<sup>29</sup> |
| 16 | Indonesia | 155.199 | Dec 31 | 217.363 | March 21 | 80 | 61.18<sup>30</sup> | 63.48<sup>27</sup> | 17.86<sup>30</sup> | 24.99<sup>27</sup> |
| 17 | Canada | 1380.218 | Dec 31 | 1629.606 | Jun 06 | 157 | 81.79 | 82.6<sup>15</sup> | 57.41 | 79.14<sup>15</sup> |
| 18 | South Africa | 357.471 | Dec 31 | 431.667 | Jun 22 | 173 | 31.83 | 35.13<sup>26</sup> | 5.79 | 7.36<sup>26</sup> |
| 19 | Egypt | No data |  | 109.415 | May 01 |  | 33.6<sup>5</sup> | 38.15<sup>24</sup> | 5.03<sup>5</sup> | 13.71<sup>24</sup> |
| 20 | Israel | 3637.259 | Dec 31 | 5573.137 | Jun 22 | 173 | 65.09 | 65.19<sup>21</sup> | 56.45 | 61.03<sup>21</sup> |
| 21 | Nigeria | 17.916 | Dec 26 | 24.74 | Jun 22 | 178 | 9.6<sup>4</sup> | 31.94<sup>16</sup> | 0.54<sup>4</sup> | 5.63<sup>16</sup> |
| 22 | Australia | 2120.241 | Dec 31 | 2830.525 | Jun 22 | 173 | 82.7<sup>3</sup> | 82.7<sup>18</sup> | 62.19 | 75.6<sup>15</sup> |
| 23 | New Zealand | 1087.221 | Dec 31 | 1416.09 | Jun 23 | 174 | 79.34 | 80.66<sup>19</sup> | 52.95 | 68.81<sup>19</sup> |
| 24 | Vietnam | 765.759 | Dec 30 | 880.538 | Jun 20 | 172 | 81.67<sup>7</sup> | 87.55<sup>28</sup> | 57.03<sup>1</sup> | 59.05<sup>28</sup> |
| 25 | European Union | No data |  | No data |  |  | 72.56 | 72.86 | 52.15 | 62.09 |
| 26 | Europe | No data |  | No data |  |  | 65.26 | 66.21 | 40.44 | 48.22 |
| 27 | Africa | No data |  | No data |  |  | 17.89 | 31.36 | 2.1 | 6.4 |
| 28 | Asia | No data |  | No data |  |  | 70.16 | 73.22 | 28.35 | 38.23 |
| 29 | North America | No data |  | No data |  |  | 63.72 | 65.7 | 37.23 | 41.9 |
| 30 | South America | No data |  | No data |  |  | 74.75 | 77.12 | 47.5 | 58.31 |
| 31 | High income countries | No data |  | No data |  |  | 73.18 | 74.3 | 51.83 | 66.37 |
| 32 | Upper middle income countries | No data |  | No data |  |  | 77.15 | 78.74 | 45.55 | 49.97 |
| 33 | Lower middle income countries | No data |  | No data |  |  | 53.28 | 59.36 | 9.17 | 19.35 |
| 34 | The world | No data |  | No data |  |  | 60.07 | 64.66 | 26.58 | 34.93 |

Figures corresponding to different days in 2022:
May: <sup>1</sup> – 18;
June: <sup>2</sup> – 1; <sup>3</sup> – 17; <sup>4</sup> – 19; <sup>5</sup> – 29; <sup>6</sup> – 30; <sup>30</sup> – 21;
July: <sup>7</sup> – 7;
September: <sup>8</sup> – 1; <sup>9</sup> – 4; <sup>10</sup> – 11;
October: <sup>11</sup> – 7;
November: <sup>12</sup> – 18; <sup>13</sup> – 22;
December: <sup>14</sup> – 12.
Figures corresponding to different days in 2023:

To calculate the averaged daily numbers of cases *DCC* and deaths *DDC* per million in 2022 and 2023 we will use simple formulas:

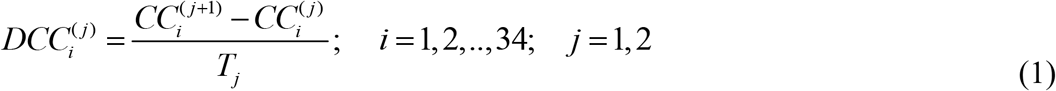

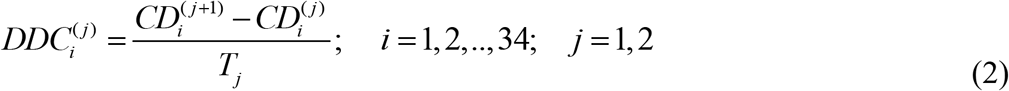

where *T*_*1*_ =365 and *T*_*2*_ =252. The *CFR* values corresponding to 2022 and 2023 can be calculated as follows:

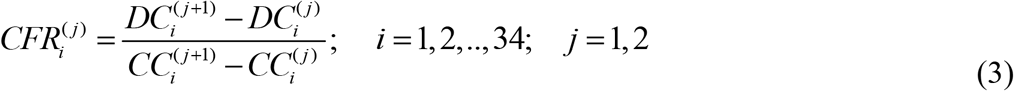

The total average *CFR*_*i*_^*\**^ levels (during the entire period of the COVID-19 pandemic) can be calculated for every country and region:

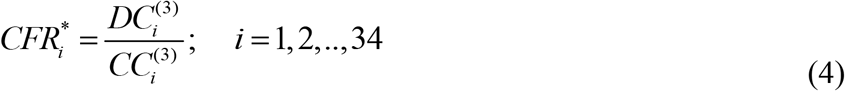

To estimate the average daily numbers of tests per thousand, we will use the formula:

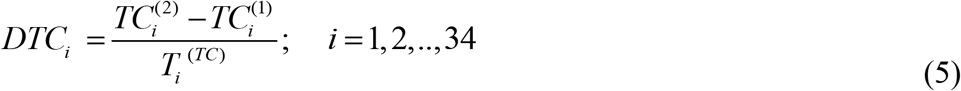

Durations of corresponding periods in days *T*_*i*_^(*TC*)^ are listed in Table 2. Unfortunately, the testing data is not available for many countries and regions.

We will use the linear regression to calculate the regression coefficients *r* and the optimal values of parameters *a* and *b* for corresponding best fitting straight lines, [47]:

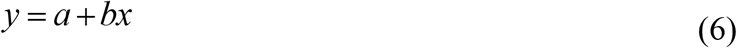

where explanatory variables *x* will be A, DTC, VC, and BC and dependent variables *y* will be DCC, DDC, CFR, DTC, VC, and BC.

We will use also the F-test for the null hypothesis that says that the proposed linear relationship (6) fits the data sets. The experimental values of the Fisher function can be calculated using the formula:

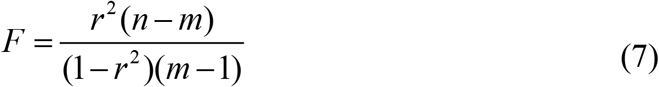

where *n* is the number of observations (number of countries and regions taken for statistical analysis); *m*=2 is the number of parameters in the regression equation, [47]. The corresponding experimental values *F* have to be compared with the critical values *F*_*C*_ (*k*_1_, *k*_2_ ) of the Fisher function at a desired significance or confidence level ( *k*_1_ = *m* −1, *k*_2_ = *n* − *m*, see, e.g., [48]). If *F* / *F*_*C*_ (*k*_1_, *k*_2_ ) < 1, the correlation is not supported by the results of observations. The highest values of *F* / *F*_*C*_ (*k*_1_, *k*_2_ ) correspond to the most reliable correlation.

We will use also non-linear regression:

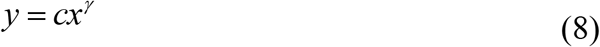

which can be reduced to the linear one with the use of new variables, [19]:

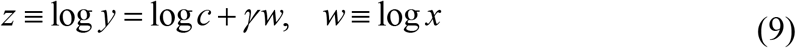

## Results

The results of calculations with the use of formulas (1)-(5) are listed in Table 3 and shown in Figures 1-3 versus median age, testing level *DTC*, percentage of fully vaccinated persons 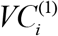 (for 2022) and 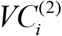 (for 2023), and numbers of boosters per hundred 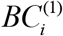 (for 2022) and 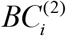 (for 2023). The averaged daily numbers of cases decreased drastically in 2023 in comparison with corresponding values in 2022 (compare *DCC* ^*(2)*^ and *DCC* ^*(1)*^ figures in Table 3 or “triangles” and “circles” in Fig. 1; the only exception is Nigeria). The global average daily number of cases has diminished 7.4 times in 2023 (see the last row of Table 3). Turkey has stopped to show new cases in 2023. USA, China, Japan do not report any COVID-19 cases and related deaths since May, 15, 2023, [1].

**Table 3.** The results of calculations of average daily characteristics and the total average CFR levels with the use of eqs. (1)-(5).

| No.,<br>I | Country<br>or region | Average daily<br>numbers of<br>COVID-19 cases<br>per million, $DCC^{(j)}$ | | Average daily numbers<br>of deaths per million,<br>$DDC^{(j)}$ | | Case Fatality Risks | | | Average<br>daily<br>numbers<br>of tests<br>per<br>thousand,<br>$DTC_i$ |
| --- | --- | --- | --- | --- | --- | --- | --- | --- | --- |
| | | 2022 | 2023 | 2022 | 2023 | 2022,<br>$CFR^{(1)}$ | 2023,<br>$CFR^{(2)}$ | Total,<br>$CFR^*$ | |
| 1 | USA | 371.55 | 47.22 | 2.1332 | 0.5243 | 0.00574 | 0.0111 | 0.0109 | 3.27 |
| 2 | Taiwan | 1012.53 | 231.08 | 1.6515 | 0.6223 | 0.00163 | 0.00269 | 0.00186 | 1.95 |
| 3 | Hong<br>Kong | 955.93 | 152.96 | 4.2416 | 1.2654 | 0.00444 | 0.00827 | 0.00487 | 13.74 |
| 4 | India | 19.02 | 0.89 | 0.0959 | 0.00371 | 0.00504 | 0.00415 | 0.0118 | 0.75 |
| 5 | France | 1241.26 | 61.89 | 1.5898 | 0.3879 | 0.00128 | 0.00627 | 0.00431 | 7.21 |
| 6 | Germany | 993.35 | 56.92 | 1.5789 | 0.4399 | 0.00159 | 0.00772 | 0.00455 | 2.77 |
| 7 | Brazil | 178.63 | 26.11 | 0.9533 | 0.2013 | 0.00534 | 0.00771 | 0.0187 | 0.32 |
| 8 | South<br>Korea | 1502.90 | 422.18 | 1.4089 | 0.2804 | 0.000937 | 0.000664 | 0.00104 | 6.39 |
| 9 | Japan | 604.99 | 150.42 | 0.8647 | 0.5500 | 0.00142 | 0.00365 | 0.00221 | 1.18 |
| 10 | Italy | 890.40 | 55.14 | 2.2064 | 0.4446 | 0.00248 | 0.00806 | 0.00736 | 8.27 |
| 11 | UK | 436.38 | 30.30 | 1.5950 | 0.7754 | 0.00366 | 0.02559 | 0.00928 | 11.76 |
| 12 | Turkey | 242.79 | 0 | 0.6201 | 0 | 0.00255 | - | 0.00596 | 3.45 |
| 13 | Mexico | 69.55 | 12.65 | 0.5947 | 0.09527 | 0.00855 | 0.00753 | 0.0436 | 0.16 |
| 14 | Peru | 174.34 | 7.52 | 1.2547 | 0.3831 | 0.00720 | 0.05093 | 0.0490 | 2.24 |
| 15 | Iran | 42.34 | 2.41 | 0.4056 | 0.07502 | 0.00958 | 0.03119 | 0.0192 | 0.77 |
| 16 | Indonesia | 24.43 | 1.35 | 0.1643 | 0.01881 | 0.00672 | 0.01396 | 0.0238 | 0.78 |
| 17 | Canada | 170.30 | 21.13 | 1.3404 | 0.4516 | 0.00787 | 0.02138 | 0.0113 | 1.59 |
| 18 | South Africa | 27.53 | 1.59 | 0.5264 | 0.00179 | 0.01911 | 0.00113 | 0.0252 | 0.43 |
| 19 | Egypt | 3.22 | 0.0175 | 0.0759 | 0.001004 | 0.02351 | 0.05730 | 0.0481 | - |
| 20 | Israel | 945.19 | 81.64 | 0.9902 | 0.4161 | 0.00105 | 0.00510 | 0.00262 | 11.19 |
| 21 | Nigeria | 0.31 | 0.365 | 0.001567 | 0 | 0.00501 | 0 | 0.0110 | 0.0383 |
| 22 | Australia | 1090.44 | 122.21 | 1.6089 | 0.8726 | 0.00148 | 0.00713 | 0.00203 | 4.11 |
| 23 | New Zealand | 1079.82 | 245.18 | 1.2047 | 0.7370 | 0.00112 | 0.00301 | 0.00139 | 1.89 |
| 24 | Vietnam | 273.28 | 3.96 | 0.3011 | 0.0008095 | 0.00110 | 0.000204 | 0.00371 | 0.67 |
| 25 | European Union | 763.24 | 42.94 | 1.7408 | 0.3750 | 0.00228 | 0.00873 | 0.00676 | - |
| 26 | Europe | 573.43 | 37.18 | 1.6369 | 0.3499 | 0.00285 | 0.00941 | 0.00833 | - |
| 27 | Africa | 6.05 | 0.358 | 0.05689 | 0.001425 | 0.00941 | 0.00398 | 0.0197 | - |
| 28 | Asia | 110.21 | 21.86 | 0.1613 | 0.08303 | 0.00146 | 0.00380 | 0.00543 | - |
| 29 | North America | 252.82 | 32.77 | 1.4811 | 0.3544 | 0.00586 | 0.01082 | 0.0129 | - |
| 30 | South America | 168.68 | 18.68 | 0.9360 | 0.1311 | 0.00555 | 0.00702 | 0.0197 | - |
| 31 | High income countries | 590.15 | 69.68 | 1.5920 | 0.4223 | 0.00270 | 0.00606 | 0.00682 | - |
| 32 | Upper middle income countries | 154.51 | 28.71 | 0.3990 | 0.1541 | 0.00258 | 0.00537 | 0.0109 | - |
| 33 | Lower middle income countries | 24.61 | 1.14 | 0.1147 | 0.009516 | 0.00466 | 0.00837 | 0.0137 | - |
| 34 | The world | 152.54 | 20.54 | 0.4277 | 0.1193 | 0.00280 | 0.00581 | 0.00903 | - |

**Fig. 1.**
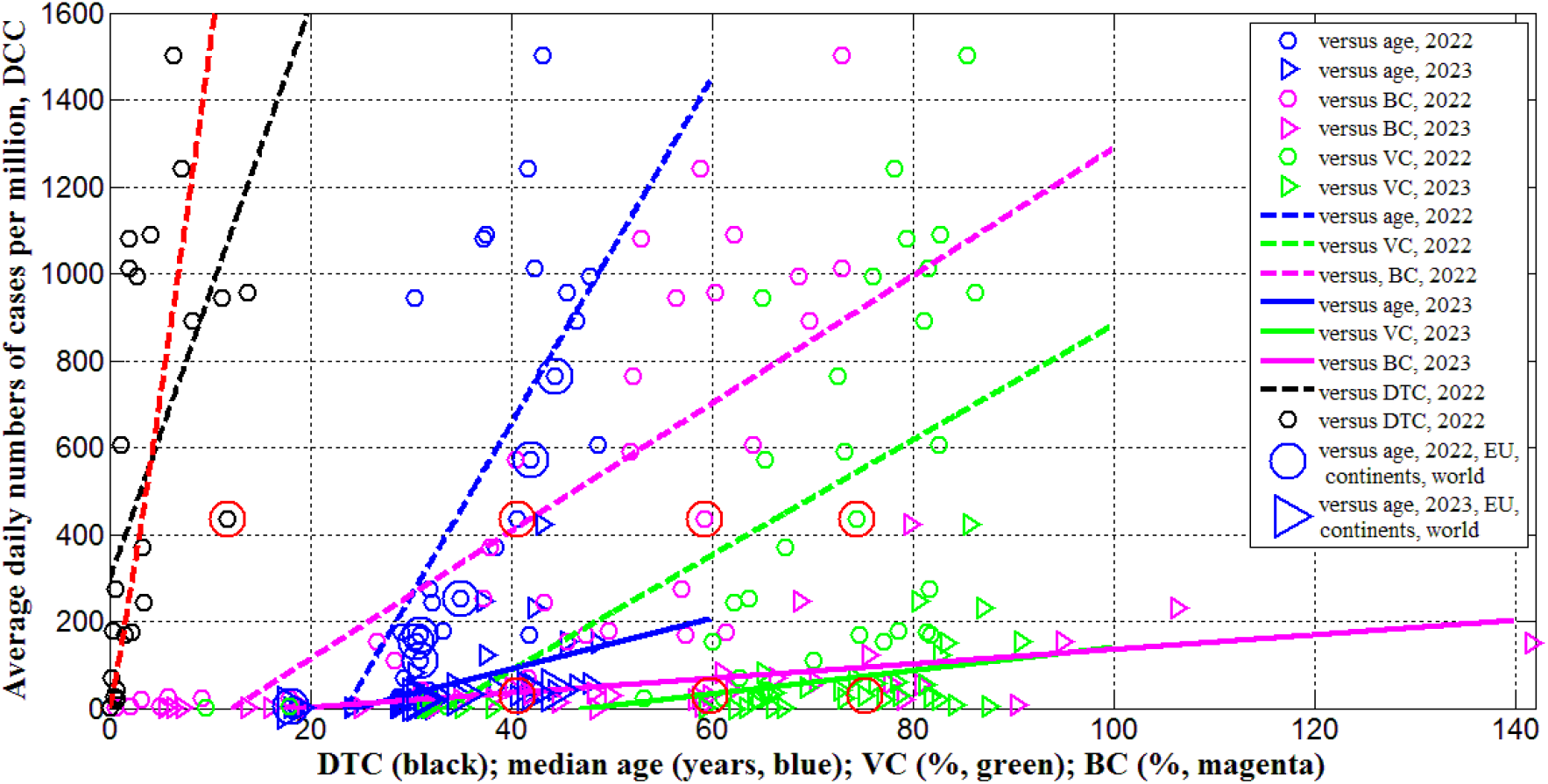
Averaged daily numbers of COVID-19 cases per million in 2022 (“circles”) and 2023 (“triangles”) versus median age (blue) and levels of vaccinations (green), boosters (magenta) and testing (black). Best fitting lines are solid for 2023 and dashed for 2022. The UK data are located in red circles. The values corresponding to EU, continents and the world are duplicated by lager markers. The red curve represents the results of non-linear correlation (eq. (10)).

In 2023 the averaged daily numbers of deaths significantly decreased in all countries an regions (compare *DDC* ^*(2)*^ and *DDC* ^*(1)*^ values in Table 3 or “triangles” and “circles” in Fig. 2), yielding 3.6 times decrease in global *DDC* figures (see the last row of Table 3). Seasonal global influenza mortality is between 294 and 518 thousand in the period from 2002 to 2011, [49]. After dividing the presented figures over the world population 8060.5 million [45] and 365 days, the corresponding averaged daily number of deaths per million *DDC*_*(infl)*_ will range between 0.1 and 0.18. The global value of 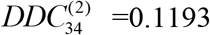 is comparable with the influenza mortality, but in 2023 in many countries (including the UK) the corresponding *DDC* ^*(2)*^ values are much higher than *DDC*_*(infl)*_ (see Table 3).

**Fig. 2.**
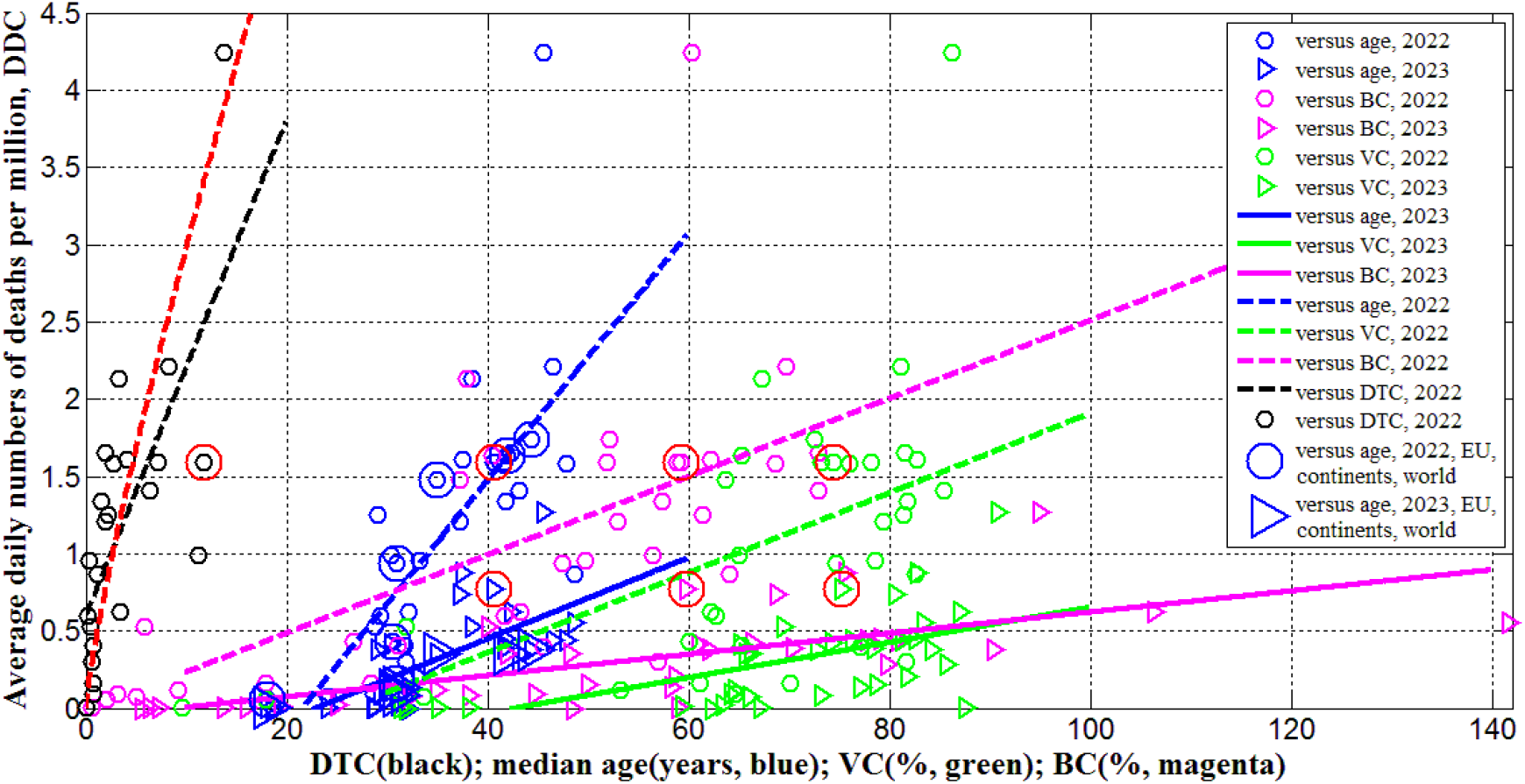
Averaged daily numbers of COVID-19 related deaths per million in 2022 (“circles”) and 2023 (“triangles”) versus median age (blue) and levels of vaccinations (green), boosters (magenta) and testing (black). Best fitting lines are solid for 2023 and dashed for 2022. The UK data are located in red circles. The values corresponding to EU, continents and the world are duplicated by lager markers. The red curve represents the results of non-linear correlation (eq. (11)).

The global case fatality risk in 2023 is approximately twice higher than in 2022 despite of the increase in percentages of fully vaccinated people and boosters (see the last rows of Tables 2 and 3). In 2023 the *CFR* values were lower only in India, South Korea, Mexico, South Africa, Nigeria, Vietnam and Africa (compare corresponding columns in Table 3). There are countries with drastic growth of the *CFR* values in 2023 in comparison with 2022 (for example, almost 7 times for the UK and Peru). In 2023 only Egypt, Peru and Iran have higher case fatality risks than one in the UK (see Table 3; the markers corresponding to the UK are placed inside red circles in Figs. 1-3).

There are countries with traditional high levels of *CFR*. To smooth temporarily fluctuations, the total average *CFR*_*i*_*\** values (during the entire period of the COVID-19 pandemic) were calculated with the use of eq. (4), listed in Table 3 and shown in Fig. 3 by blue “stars”. The value *CFR*_*11*_*\** corresponding to the UK is only slightly higher than the global one. Many countries (e.g., the US, India, Mexico, Peru, Iran, Indonesia, Canada, South Africa, Egypt, Nigeria) have higher *CFR*_*i*_*\** values.

**Fig. 3.**
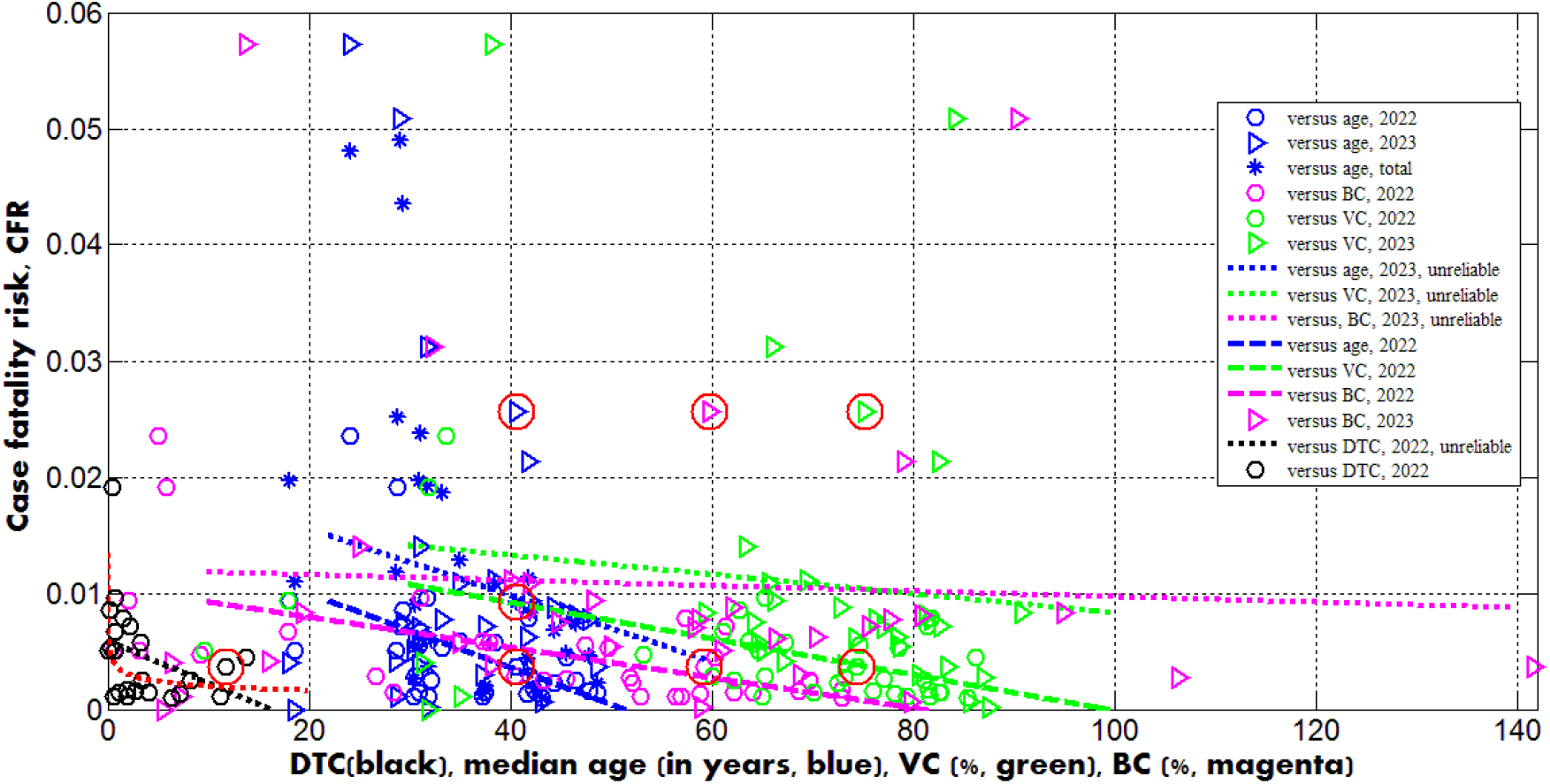
Case fatality risks in 2022 (“circles”) and 2023 (“triangles”) versus median age (blue) and levels of vaccinations (green), boosters (magenta) and testing (black). Best fitting lines are solid for 2023 and dashed for 2022. The dotted lines correspond to the correlations that are not supported at the significance level 0.01. The UK data are located in red circles. The red curve represents the results of non-linear correlation (eq. (12)).

To remove the influence of the seasonal factors in the UK, let us calculate the values of 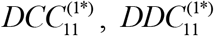 and 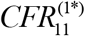 for the period January 1, 2022 – May 19, 2022 and the same values 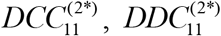 and 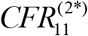 and for the period January 1, 2023 – May 19, 2023 with the use of eqs. (1)-(4) and JHU datasets:

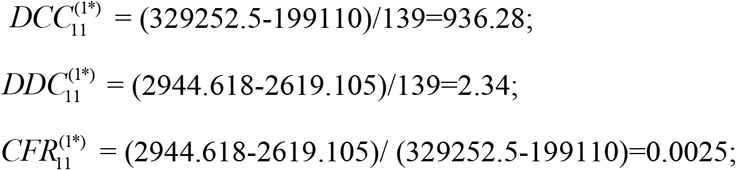

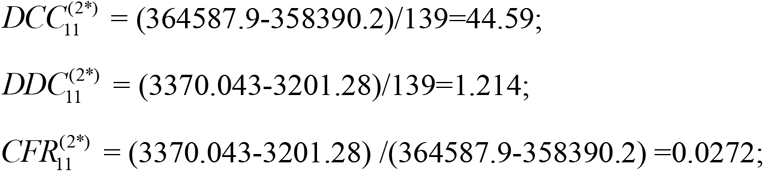

In 2023 we can see huge decrease in the number of cases and moderate diminishing in the number of death. As the result, the case fatality risk in the beginning of 2023 exceeded the 2022 figure around 11 times. The reason could be explained by the fact that testing and reporting most cases we stopped in UK since April 2022. The only people who get recorded as cases are those that are tested in hospital, and even there not everyone with respiratory infections gets tested. The death data may include all those where COVID-19 is listed on the death certificate, which might even include those that have not tested positive but where the doctors suspect COVID-19.

It must be noted, the growth of *CFR* in the UK occurred in the period of increasing the vaccination and booster levels, [2]:

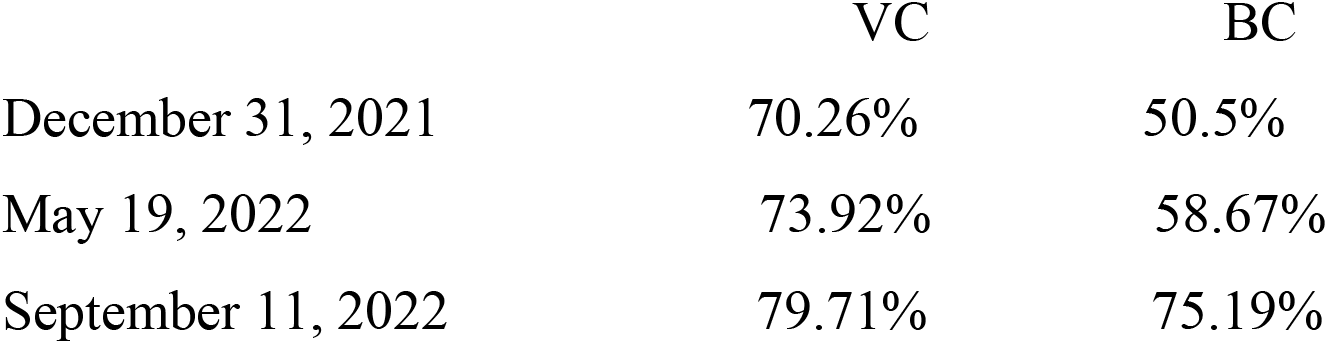

To investigate possible correlations between *DCC, DDC* and *CFR* values and explanatory variables *A, DTC, VC*^*(1)*^, *VC*^*(2)*^, *BC*^*(1)*^, and *BC*^*(2)*^, the linear regression (6) and Fisher test were used. The results of calculations of optimal values of parameters *a* and *b*, correlation coefficients and experimental values of the Fisher function *F* (eq. (7)) are listed in Table 4. The *F* values were compared with the critical ones *F*_*C*_ (1, *n* − 2) at the confidence level 0.01. The numbers of observations are different for different correlations due to the absence of data for some countries and regions.

**Table 4.** Optimal values of parameters in eq. (1), correlation coefficients and the results of Fisher test applications.

| Number | Variable $x$ | Number of observations $n$ | Correlation coefficient $R$ | Optimal values of parameter $a$ in eq. (6) | Optimal values of parameter $b$ in eq. (6) | Experimental value of the Fisher function $F$, eq. (7), $m=2$ | Critical value of Fisher function $F_c(l, n-2)$ for the confidence level 0.01, [29] | $F/F_c$ |
| --- | --- | --- | --- | --- | --- | --- | --- | --- |
| **Correlations for the averaged daily numbers of COVID-19 cases in 2022, $DCC^{(1)}$** |  |  |  |  |  |  |  |  |
| 1 | versus age | 31 | 0.7108 | -937.43 | 39.767 | 29.616 | 7.77 | 3.81 |
| 2 | versus $VC^{(1)}$ | 34 | 0.56202 | -444.10 | 13.252 | 14.775 | 7.74 | 1.91 |
| 3 | versus $BC^{(1)}$ | 34 | 0.7459 | -181.28 | 14.687 | 40.136 | 7.74 | 5.19 |
| 4 | versus $DTC$ | 23 | 0.5613 | 292.96 | 66.052 | 9.66 | 7.85 | 1.2 |
| **Correlations for the averaged daily numbers of COVID-19 cases in 2023, $DCC^{(2)}$** |  |  |  |  |  |  |  |  |
| 5 | versus age | 31 | 0.5009 | -143.583 | 5.8231 | 9.711 | 7.77 | 1.25 |
| 6 | versus<br>$VC^{(2)}$ | 34 | 0.4696 | -128.811 | 2.6796 | 9.051 | 7.74 | 1.17 |
| 7 | versus<br>$BC^{(2)}$ | 34 | 0.5674 | -32.3957 | 1.6692 | 15.195 | 7.74 | 1.96 |
| **Correlations for the averaged daily numbers of deaths in 2022, $DDC^{(1)}$** |  |  |  |  |  |  |  |  |
| 8 | versus<br>age | 31 | 0.7324 | -1.7244 | 0.07993 | 33.553 | 7.77 | 4.32 |
| 9 | versus<br>$VC^{(1)}$ | 34 | 0.5553 | -0.6756 | 0.02582 | 14.264 | 7.74 | 1.84 |
| 10 | versus<br>$BC^{(1)}$ | 34 | 0.6513 | -0.02176 | 0.02529 | 23.579 | 7.74 | 3.05 |
| 11 | versus<br>$DTC$ | 23 | 0.7138 | 0.59477 | 0.16076 | 21.8 | 7.85 | 2.8 |
| **Correlations for the averaged daily numbers of deaths in 2023, $DDC^{(2)}$** |  |  |  |  |  |  |  |  |
| 12 | versus<br>age | 31 | 0.6793 | -0.6040 | 0.02626 | 24.843 | 7.77 | 3.20 |
| 13 | versus<br>$VC^{(2)}$ | 34 | 0.5927 | -0.4892 | 0.0114 | 17.332 | 7.74 | 2.24 |
| 14 | versus<br>$BC^{(2)}$ | 34 | 0.6903 | -0.06489 | 0.006845 | 29.131 | 7.74 | 3.76 |
| **Correlations for the case fatality risks in 2022, $CFR^{(1)}$** |  |  |  |  |  |  |  |  |
| 15 | versus<br>age | 31 | -0.5006 | 0.01631 | -0.0003177 | 9.698 | 7.77 | 1.25 |
| 16 | versus<br>$VC^{(1)}$ | 34 | -0.5901 | 0.01538 | -0.0001553 | 17.092 | 7.74 | 2.21 |
| 17 | versus<br>$BC^{(1)}$ | 34 | -0.5937 | 0.01053 | -0.0001305 | 17.423 | 7.74 | 2.25 |
| 18 | versus<br>$DTC$ | 23 | -0.3566 | 0.005909 | -0.00036525 | 3.059 | 7.85 | 0.39 |
| **Correlations for the case fatality risks in 2023, $CFR^{(2)}$** |  |  |  |  |  |  |  |  |
| 19 | versus<br>age | 30 | -0.1700 | 0.02122 | -0.00028515 | 0.834 | 7.78 | 0.11 |
| 20 | versus<br>$VC^{(2)}$ | 33 | -0.1008 | 0.01656 | -8.2800e-05 | 0.318 | 7.75 | 0.041 |
| 21 | versus<br>$BC^{(2)}$ | 33 | -0.0560 | 0.01203 | -2.36499e-05 | 0.0975 | 7.75 | 0.013 |

Since many COVID-19 patients are asymptomatic [25-30], the high testing level (*DTC* or *TC*) could help to reveal more cases and COVID-19 related deaths. This trend was supported statistically (see rows 4 and 11 in Table 4 and black lines in Figs. 1 and 2). Nevertheless, the linear regression yields unacceptable non-zero values of parameter *a*, which mean that some cases and deaths could be revealed at zero testing level. To remove this discrepancy, the non-linear approach (8)-(9) was applied for the same countries listed in Table 3 (*n*=23).

Equations (10)-(12) represent the best fitting curves (see red lines in Fig. 1-3), correlation coefficients and experimental values of the Fisher function:

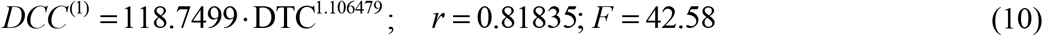

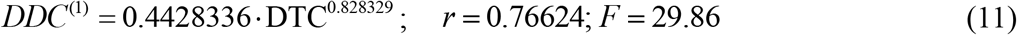

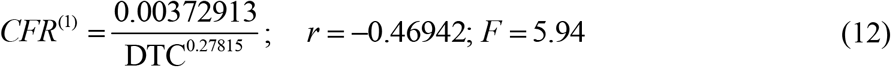

The values of *r*^*2*^ and *F* for variables *z* and *w* are higher than for *y* and *x* (compare corresponding values in (10)-(12) and in rows 4, 11 and 18 of Table 4). The relationships (10) and (11) are supported at significance level 0.001 ( *F*_*c*_ (1, 21) = 14.6 ). The similar very strong correlation between the numbers of cases and tests per capita accumulated in European and African countries as of August 1, 2022 was found in [19]:

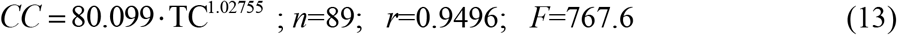

Nevertheless, 16 European countries with the highest testing level (*TC*>3000) have demonstrated no correlation between *CC* and *TC* even at significance level 0.05, [19].

Only 5 countries and territories listed in Table 2 (Hong Kong, France, Italy, the UK, and Israel) had *TC* values higher than 3,000 in 2022. The *DCC*^*(1)*^ values are rather high and vary from 436 to 1,241 in these countries (see Table 3). Nevertheless, many infectious persons were probably not detected. This is evidenced not only by the higher numbers of cases per capita in South Korea (*DCC*_*8*_^*(1)*^=1,502.9, Table 3) at lower testing level (compare corresponding *DTC* values in Table 3), but also by the results of total testing in some countries and institutions, which revealed many previously unregistered COVID-19 patients [24-26]. Taking the maximum of *DCC*_*8*_^*(j)*^ values (corresponding to South Korea) as estimations real number of cases per capita in 2022 and 2023, we can calculate the visibility coefficients

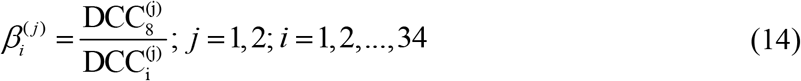

as the ratios of real and registered numbers of cases (similar relationship can be obtained using the accumulated numbers of cases per capita, [19]). For example, figures corresponding to the UK are 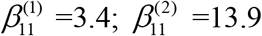; Europe - 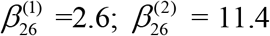, and Africa - 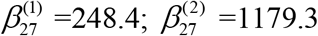.

An experimental estimation of the visibility coefficient can be obtained from the results of total testing in Slovakia (89.5% of population was tested on October 31-November 7, 2020 and a number of previously undetected cases, equal to about 1.63% of the population was revealed, [24, 25]). Since the number of detected cases in Slovakia was approximately 1% of population [2], we can estimate the visibility coefficient *β* ≈ 2.63 for that period. As of September 10, 2023 the ratio of CC values for South Korea and Slovakia (667,207.1/330,868.413, [2]) yields the visibility coefficient 2.02. The results of a random testing in two kindergartens and two schools in Chmelnytskii (Ukraine) revealed the value of visibility coefficient 3.9 in December 2020, [26].

The generalized SIR models and algorithms of their parameter identification [27, 29, 50] allowed theoretical estimating of the visibility coefficients. In particular, values from 3.7 to 20.4 were obtained for Ukraine [15, 27] and 5.4 for Qatar [28] in different periods of the COVID-19 pandemic. The lack of appropriate testing did not allowed detecting the first SARS-CoV-2 cases, which probably appeared long before December 2019 [51]. In particular, theoretical estimates give the date of the appearance of the first case at the beginning of August 2019, [50].

Dependence (12) can be accepted at significance level 0.05 ( *F*_*c*_ (1, 21) = 4.43 ; a similar equation can be obtained by dividing (11) over (10)) and shows that the case fatality risk increases with diminishing of the testing level even in period of the high interest in the SARS-CoV-2 infection (as it was in 2022). In 2023, when the people paid attention to severe cases only and maked tests correspondingly, *CFR* values can increase drastically. Therefore, one should probably not be afraid of a significant increase of the case fatality risk in the UK in 2023. Of much greater concern is the fact that COVID-19 mortality in this country ( 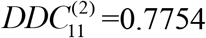, see Table 3) is still at least 4 times higher than the global value caused by seasonal flu, [49].

Equations (10) and (13) may give the illusion that the low number of cases per capita in Africa is due only to the low testing level typical for low-income countries (see [19]). The visibility coefficients and another characteristic - the ratio of the number of tests to the number of cases *DTS*-will allow us to understand the situation and draw the right conclusions. High *DTS* values mean that many persons surrounding the detected infectious patient (e.g., family members, colleagues, neighbors) were tested and isolated (this causes a decrease in the number of new infections, i.e. *DCC*). For example, very high tests per case ratios (*DTS* >100) in Hong Kong in 2020 and 2021 allowed controlling the COVID-19 epidemic completely [20] (the smoothed daily numbers of new cases per million did not exceed 20, [2]). After January 18, 2022, the daily numbers of new cases started to increase, but the daily numbers of tests remained almost constant yielding drastically diminishing of the daily tests per case ratio [20] and very high *DCC* values in February-March 2022, [2].

It follows from (10) that the averaged daily test per case ratio:

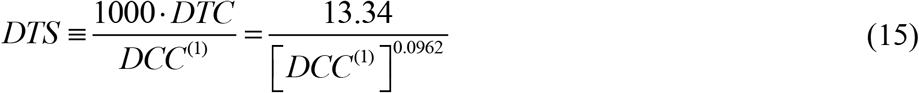

increases for countries with low *DCC* figures (in particular, for African ones, see Table 3). The similar relationship follows from (13) for the accumulated characteristic *TS=1000*TC/CC*. For example, *DTS* values (calculated using the information available in Table 3) are equal to 26. 9 (the UK); 39.4 (India); 123.5 (Nigeria); 4.3 (South Korea); 1.95 (Japan) and demonstrate that the probability to miss an infectious person due to the lack of tests is much higher in Japan or South Korea than in Nigeria or India. During the severe pandemic wave in Japan in summer 2022, the daily numbers tests probably were not enough to confirm COVID-19 in patients with symptoms [29].

Therefore, the reason for the low number of registered cases per capita in Africa or in India should not be found in insufficient testing, but in large values of the visibility coefficients (14), which attribute to large numbers of asymptomatic infections. Since the severity of SARS-CoV-2 infection increases for older patients [16, 31] and almost half of the infected children can be asymptomatic [30], the regions with older population are expected to have much higher accumulated numbers of cases per capita, [31]. It was shown that one-year increment in the median age yields 12,000-18,000 increase in *CC* values [21]. Rows 1 and 5 in Table 4 and blue lines in Fig. 1 illustrate the same trend for *DCC* values. One-year increment in the median age increased *DCC* values by 39.8 in 2022 and by 5.8 in 2023.

The stronger correlations and same trends were obtained for the averaged daily numbers of deaths per capita *DDC* versus median age of population *A* (see rows 8 and 12 in Table 4 and blue lines in Fig. 2). One-year increment in the median age increases the *DDC* values by 0.0799 in 2022 and by 0.0263 in 2023. The characteristics calculated for large regions (EU, continents and the world) are very close to the best fitting blue lines (see large markers in Figs. 1 and 2). We can conclude that the young age of Africa (*A*_*27*_ =18, see Table 1) is the main reason of very low numbers of cases and death per capita registered on this continent.

Opposite and much weaker age trends we can see for the case fatality risks (lines 15 and 19 in Table 4). The decrease of *CFR* values with increase of the age (supported only by the 2022 dataset) looks unexpected (especially taking into account the fact that in 2020 younger populations had less clinical cases per capita [31]). Probably, the reasons are higher vaccination levels and better medical treatment in the reach countries with high median age.

The numbers of cases and deaths per capita increase with increasing the percentages of fully vaccinated people and boosters (see rows 2, 3, 6, 7, 9, 10, 13, and 14 in Table 4 and green and magenta best fitting lines in Figs. 1 and 2). Re-infections in vaccinated persons are common [52, 53], but a very clear uprising trend with increasing *VC* and *BC* values is unexpected despite the similar result for smoothed daily numbers of cases reported in [17] (JHU datasets with 7-days-smoothing corresponding to September 1, 2021 and February 1, 2023 were used for statistical analysis). Obtained trends could be a result of age influence; since the most vaccinated countries have higher *A*_*i*_ values (see Tables 1 and 2). We will discuss this correlation in the next Section. Another reason could be the introduction of special passports that removed restrictions for vaccinated persons. Many vaccinated people in countries with high *VC* and *BC* values started to visit crowded places, travel despite they can spread the infection. In many countries (in particular, in Ukraine) the vaccination procedure was associated with overcrowding in hospitals, which could contribute to the spread of the infection too.

As expected, the case fatality risks decrease with increasing the percentages of fully vaccinated people and boosters (see rows 16, 17, 20, and 21 in Table 4 and green and magenta best fitting lines in Fig. 3). Similar result was obtained in [17] with the use of JHU datasets for European and some other countries. In 2023, the decreasing trend was not supported by Fisher test. Probably, this is due to the more chaotic data. In particular, the different days correspond to 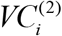 and 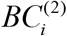 values listed in Table 2; no *CFR* value can be calculated for Turkey.

## Discussion

The explanatory variables *A, DTC, VC*^*(1)*^, *VC*^*(2)*^, *BC*^*(1)*^, and *BC*^*(2)*^, used in our analysis can be also dependent on each other. We have used the linear regression (6) and Fisher test to find correlations between *DTC, VC*^*(1)*^, *VC*^*(2)*^, *BC*^*(1)*^, and *BC*^*(2)*^ values and explanatory variable *A*. The results of calculations are listed in Table 5 and displayed in

**Table 5.** Correlations versus the median age of populations and purified levels of vaccinations. Optimal values of parameters in eq. (1), correlation coefficients and the results of Fisher test applications.

| Number | Variable<br>$y$ | Number of observations<br>$n$ | Correlation coefficient<br>$R$ | Optimal values of parameter<br>$a$<br>in eq. (1) | Optimal values of parameter<br>$b$<br>in eq. (1) | Experimental value of the Fisher function $F$,<br>eq. (3),<br>$m=2$ | Critical value of Fisher function $F_c(l, n-2)$ for the confidence level 0.01, [29] | $F/F_c$ |
| --- | --- | --- | --- | --- | --- | --- | --- | --- |
| **Correlations versus median age of population, $A$** |  |  |  |  |  |  |  |  |
| 1 | $DTC$ | 23 | 0.4389 | -4.7762 | 0.23290 | 5.011 | 7.85 | 0.64 |
| 2 | $VC^{(1)}$ | 31 | 0.7673 | 2.3288 | 1.83706 | 41.515 | 7.77 | 5.34 |
| 3 | $VC^{(2)}$ | 31 | 0.7348 | 17.5274 | 1.49084 | 34.036 | 7.77 | 4.38 |
| 4 | $BC^{(1)}$ | 31 | 0.7933 | -34.8469 | 2.21980 | 49.246 | 7.77 | 6.34 |
| 5 | $BC^{(2)}$ | 31 | 0.7810 | -51.3696 | 3.03961 | 45.339 | 7.77 | 5.84 |
| **Correlations versus “purified” numbers of fully vaccinated persons per hundred, VP** |  |  |  |  |  |  |  |  |
| 6 | $DCC^{(1)}$ | 31 | 0,03726 | 463,8161 | 1,35771 | 0,0403 | 7.77 | 0,0052 |
| 7 | $DDC^{(1)}$ | 31 | 0,00318 | 1,0919 | 0,00022628 | 0,000294 | 7.77 | 3,8e-5 |
| 8 | $CFR^{(1)}$ | 31 | -0,3236 | 0,005114 | -0,00013372 | 3,391 | 7.77 | 0,43 |
| **Correlations versus “purified” numbers of boosters per hundred, BP** |  |  |  |  |  |  |  |  |
| 9 | $DCC^{(1)}$ | 31 | 0,2969 | 463,792 | 9,75216 | 2,804 | 7.77 | 0,36 |
| 10 | $DDC^{(1)}$ | 31 | 0,0989 | 1,0919 | 0,0063360 | 0,2864 | 7.77 | 0,037 |
| 11 | $CFR^{(1)}$ | 31 | -0,3625 | 0,005117 | -0,00013502 | 4,386 | 7.77 | 0,56 |

Fig. 4. There are strong correlations between *VC*^*(1)*^, *VC*^*(2)*^, *BC*^*(1)*^, and *BC*^*(2)*^ versus median age of population *A* (see rows 2-5 in Table 5; green and magenta best fitting lines in Fig. 4). The correlation between *A* and *DTC* is supported at the confidence level 0.05 (see the first row in Table 5 and the black best fitting line in Fig. 4). The growth of the median age leads to the increase of testing level and the percentages of vaccinations and boosters. These correlations can be a result of higher incomes in aged countries and more vaccinations and boosters in older people.

**Fig. 4.**
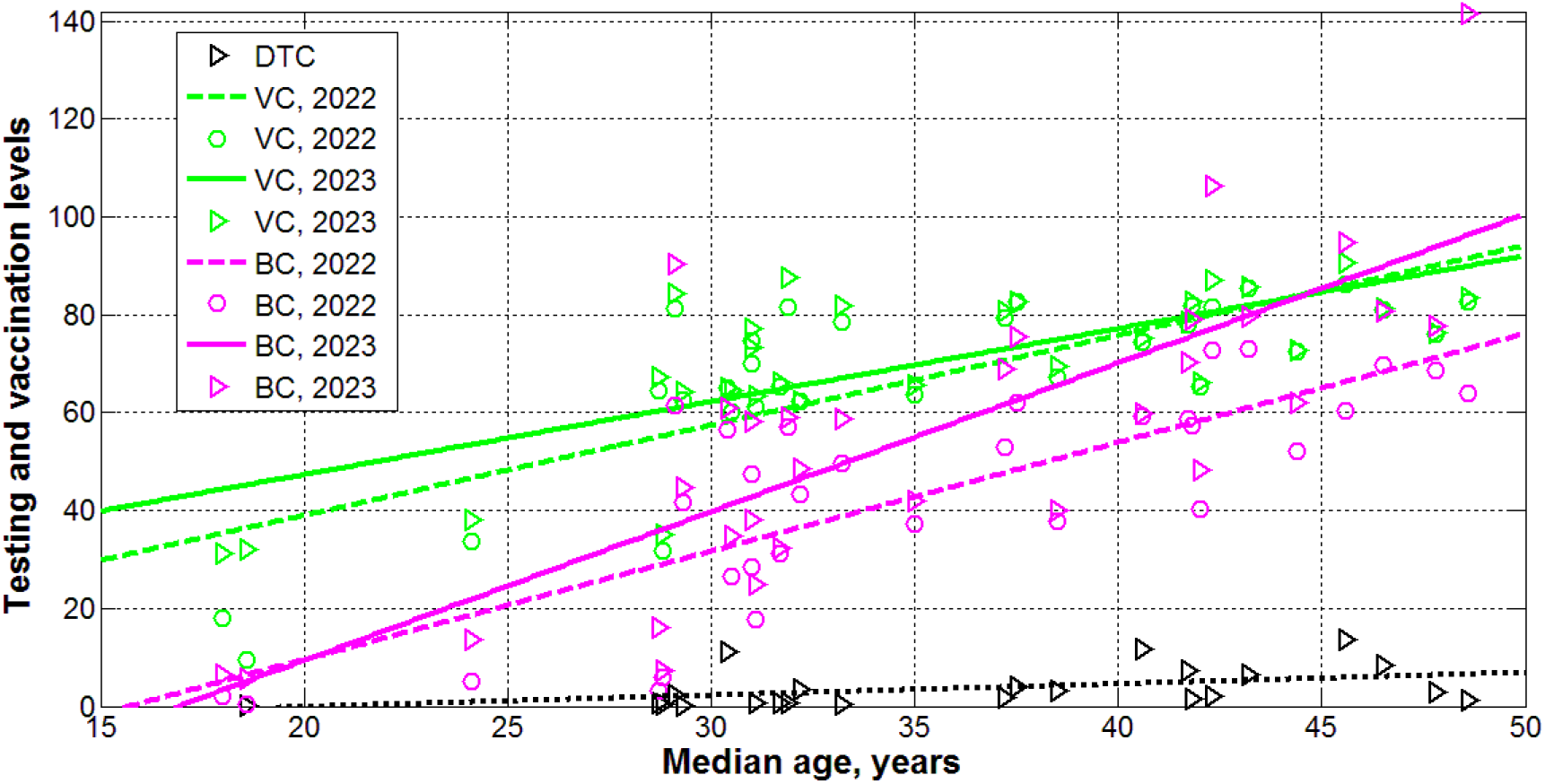
Levels of testing (black), vaccinations (green) and boosters (magenta) in 2022 (“circles”) and 2023 (“triangles”) versus median age. Best fitting lines are solid for 2023 and dashed for 2022. The dotted line corresponds to the *DTC* correlation (supported at the significance level 0.05).

Now we can explain why the numbers of cases and deaths per capita can increase with increasing the percentages of fully vaccinated people and boosters (see rows 2, 3, 6, 7, 9, 10, 13, and 14 in Table 4 and green and magenta best fitting lines in Figs. 1 and 2)? Values *VC*^*(1)*^, *VC*^*(2)*^, *BC*^*(1)*^, and *BC*^*(2)*^ are not independent and definitely increase with the age. On the other hand, *DCC* and *DDC* values also increase with growth of *A*_*i*_ (see rows 1, 5, 8, and 12 in Table 4 and blue best fitting lines in Figs. 1 and 2). To remove the influence of age in correlations between vaccinations and *DCC* and *DDC* values, let us consider the “purified” variations of *VC*^*(1)*^ and *BC*^*(1)*^ (we limited ourselves only to 2022 with more reliable statistical data):

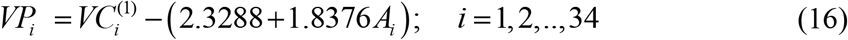

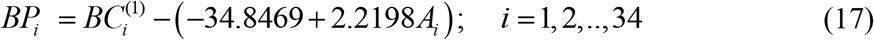

To obtain the “purified” variations the percentages of vaccinations *VP*_*i*_ and boosters *BP*_*i*_, we have excluded from variations *VC*^*(1)*^ and *BC*^*(1)*^ the values predicted by the by the best fitted lines listed in Table 5 (rows 2 and 4).

We have used the linear regression (6) and Fisher test to find correlations between *DCC, DDC* and *CFR* values and explanatory variables *VP* and *BP*. The results of calculations are listed in Table 5 (lines 6-11). No correlations were revealed at the confidence level 0.01. Thus, the vaccinations and booster themselves do not increase the numbers of cases and death per capita. As expected, the case fatality risks reduce for higher values of *VP* and *BP* (see lines 8, 11 in Table 5), but at lower confidence level than versus *VC*^*(1)*^ and *BC*^*(1)*^ (see lines 16 and 17 in Table 4).

## Conclusions

The averaged daily numbers of cases *DCC* and death *DDC* per million, case fatality risks *DDC/DCC* were calculated for 34 countries and regions with the use of John Hopkins University (JHU) datasets for numbers per capita accumulated in 2022 and 2023. Linear and non-linear approaches were used to find correlations with the averaged daily numbers of tests per thousand *DTC*, median age of population *A*, and percentages of vaccinations *VC* and boosters *BC*.

One-year increment in the median age yielded 39.8 increase in *DCC* values and 0.0799 in *DDC* figures in 2022 (in 2023 these increments are 5.8 and 0.0263, respectively). Decreasing testing level *DTC* can drastically increase the case fatality risk. *DCC* and *DDC* values grow with increasing the percentages of fully vaccinated people and boosters. Since *VC* and *BC* values definitely increase with at higher *A*, the influence of age was removed and no correlations between corrected variations of *VC* and *BC* and *DCC* and *DDC* values were obtained.

The presented analysis demonstrates that age is a pivot factor in visible (registered) part of the COVID-19 pandemic dynamics. Much younger Africa has registered less numbers of cases and death per capita due to many unregistered asymptomatic patients. Of great concern is the fact that COVID-19 mortality in 2023 in the UK is still at least 4 times higher than the global value caused by seasonal flu.

## Clarification point

No humans or human data was used during this study

## Data availability

All data generated or analyzed during this study are included in this text.

## Acknowledgement

The study was supported by the Solidarity Satellite Programme of Isaac Newton Institute for Mathematical Sciences, Cambridge, UK. The author is grateful to Professor Robin Thompson, Professor Matt Keeling, and Oleksii Rodionov for their help in providing very useful information.

